# Accurate support vector machine identification of limb-onset amyotrophic lateral sclerosis using resting-state activity of regions within executive control network

**DOI:** 10.1101/2022.08.17.22278923

**Authors:** James Meroiti

**Affiliations:** Monash University

## Abstract

Amyotrophic lateral sclerosis (ALS) is a motor neuron degenerative disorder facing diagnostic challenges due to its highly variable presentation and symptom overlap. In other neurodegenerative disorders, support vector machine (SVM) classifiers have utilized neuroimaging to address these challenges. Given functional alterations may be the earliest detectable in ALS, we aimed to uncover resting-state functional MRI (rs-fMRI) biomarkers for SVM classification. Resting-state networks derived from independent component analysis were compared between limb-onset ALS patients (n = 14) and controls (n = 12). A cluster within the executive control network (EXN) localizing predominantly to the anterior cingulate gyrus (ACG) was significantly decreased in limb-onset ALS. Activity of this cluster was able to develop a SVM with 86% sensitivity and 87% specificity on the validation dataset. These findings suggest the ACG and EXN may be important in classifying limb-onset ALS patients and could be incorporated into multi-modal SVM classifiers.

## Introduction

Amyotrophic lateral sclerosis (ALS) is a neurological disorder characterized by progressive degeneration of upper and lower motor neurons (UMNs and LMNs) resulting in severe motor dysfunction (1). Presentation of the disorder is heterogenous, with location of disease onset, rate of progression, UMN and LMN involvement, and the presence of cognitive decline varying significantly between individuals (2). Due to this heterogeneity, diagnosis of ALS is notably difficult, with approximately 50% of patients experiencing lengthy diagnostic delays (3). As such there is an unmet clinical need to uncover reliable disease biomarkers to aid diagnosis.

To address this need, several groups have developed highly sensitive ALS classifiers utilizing various MRI (4–8) and quantitative susceptibility mapping (QSM) metrics (9,10). In other neurological disorders, such as Alzheimer’s Disease (AD), multi-modal classifiers incorporating several neuroimaging modalities have been required to effectively capture the spectrum of disease presentation (11). Therefore, uncovering additional neuroimaging metrics with classification potential are necessary to improve accuracy and generalizability of multi-modal classifiers.

Resting-state functional MRI (rs-fMRI) presents a promising candidate metric, as functional alterations seem to precede structural changes and development of symptoms (12). Although resting-state network (RSN) alterations in ALS are variable, differences in several networks have been reported between ALS limb- and bulbar-onset subjects (13), which may be a contributing factor.

Therefore, this work aimed to investigate RSNs in a purely limb-onset ALS cohort, to reduce intra-group heterogeneity, and determine if activity within these networks could develop an effective support vector machine (SVM) classifier. It was hypothesized that activity across entire RSNs would not be effective in distinguishing limb-onset ALS subjects, however regions of networks showing differential activity between patients and controls would be develop a highly sensitive and specific classifier (>80%). Successfully identifying regions capable of such would enable incorporation of RSN activity in these areas into multi-modal classifiers improving performance and generalizability, thus nearing use of neuroimaging classifiers for clinical diagnosis.

## Methods

### Participants

This study investigated fourteen participants (n = 14, two female) with clinically possible, probably or definite limb-onset ALS as defined by the El Escorial diagnostic criteria (14) and eleven (n = 11, two female) gender matched control subjects who were screened for neurological disorders (Table 1). All participants recruited were over 18 years of age and able to give informed consent (Project Number: CF14/3968-2014002057).

**Table 1.** Participant demographics. Data values shown are mean values ± SEM. ALSFRS-R = ALS Functional Rating Scale-Revised, ALS-FTD-Q = ALS-Frontotemporal Dementia Questionnaire, ECAS = Edinburgh Cognitive & Behavioural ALS Screen.

|  | ALS | Control |
| --- | --- | --- |
| Number of subjects | 14 (two female) | 11 (two female) |
| Age (years) | 54.72 ± 3.49 | 51.91 ± 3.49 |
| Time since disease onset (years) | 3.25 ± 1.03 | N/A |

### MRI data acquisition

MR images were acquired using a 3-Tesla Skyra scanner (Siemens, Erlangen, Germany) at Monash Biomedical Imaging, Melbourne, Australia. Participants were instructed to fixate their gaze on a cross in the monitor for the duration of the scan. Functional MRI data consisted of 400 volumes of an echo planar sequence (TR = 801, TE = 21 ms, axial slices = 45, matrix= 64 × 64, voxel resolution = 3 × 3 × 3 mm, slice thickness 3 mm, interslice gap = 0mm, 50° flip angle). T1-weighted high-resolution MPRAGE images were acquired for co-registration of the functional datasets (TR = 2300, TE = 2.07 ms, voxel resolution 1 × 1 × 1 mm, field of view = 240 × 256 mm, 9° flip angle, 192 slices per volume).

### MRI pre-processing

Pre-processing steps were carried using FEAT (Version 6.00) a part of the FSL toolbox (www.fmrib.ox.ac.uk/fsl). The following pre-processing steps were applied to functional data: (i) motion correction through multi-resolution rigid body co-registration using MCFLIRT (15); (ii) removal of nonbrain structures from volumes using BET (16); (iii) correction for slice timing; (iv) spatial smoothing with a 5 mm full-width half-maximum Gaussian kernel to reduce individual variability (17); (v) bias removal by normalizing each volume to its grand-mean intensity; (vi) correction for distortion with gradient-echo field map; and (vii) highpass temporal filtering with 100 second cut-off. Structural images were brain-extracted using BET and visually inspected. Registration of functional data to T1-weighted and brain extracted Montreal Neurological Institute (MNI152) 1 mm standard images was carried out using Boundary-Based Registration and 12-DOF affine transformation using FLIRT (15,18) followed by FNIRT nonlinear registration (19,20).

### Independent component analysis

MELODIC (version 3.15) part of FSL was used to perform probabilistic independent component analysis (PICA) (21) for each subject where model order was estimated using the Laplace approximation (range 35-76) (22). FSL AROMA was used to automatically identify and remove motion related components from the data (23,24) with the remaining components transformed to MNI152 1mm standard space (25).

Control group data was temporally concatenated using group PICA with model order of 25 (21). For each resulting component, the component with the highest cross-correlation value to each RSN was determined to represent that network (26). These group spatial maps were then regressed into each individual dataset using dual regression to derive individual spatial maps and corresponding time courses (27,28). Each voxel contains a β-coefficient representing the estimated contribution of that voxel to the regression model (27).

### Comparison of resting-state networks

Between-group comparisons of individual spatial maps were performed using non-parametric permutation tests using FSL’s Randomise with 5,000 permutations (29). Results were corrected for multiple comparisons using threshold-free cluster enhancement (TFCE) (30) with P values less than 0.05 considered to be statistically significant. Statistically significant clusters were masked for non-brain regions. To calculate the number of voxels of clusters belonging to anatomical regions, binary masks of each anatomical region within each cluster were created and subsequently used to mask the clusters to produce significant voxels parcellated by each anatomical region, with the number of voxels within these parcellated maps calculated.

### Support vector machine classification of limb-onset ALS

To classify limb-onset ALS patients from controls, a mask of significant clusters from between-group analysis was created for each subject from individual spatial maps, with ß-coefficients of these regions averaged. Mean ß-coefficients of significant clusters as well as each of the ten RSNs, and class (0 = control, 1 = ALS patient) for each subject was used as input to train a SVM classifier using the scikit-learn library (https://scikit-learn.org/stable/) implemented in Python 3.8.5.

Data was randomly divided into training (70%) and validation datasets (30%) and stratified by group to ensure an equal division of class in each dataset. Each SVM was trained using a linear kernel, C-parameter value of 1, and default value of γ where γ = where is the training data ß-coefficient variance. After training, the SVM returned the probability of each subject in the validation dataset belonging to the limb-onset ALS group. Ten-fold cross-validation was performed using random splits of the data. Resulting probabilities and actual class from each of the ten splits were concatenated to generate receiver-operator-characteristic (ROC) curves and area-under-curve (AUC) values.

## Results

Figure 1 shows the ten group ICA components most closely resembling each of the RSNs (26).

**Figure 1.**
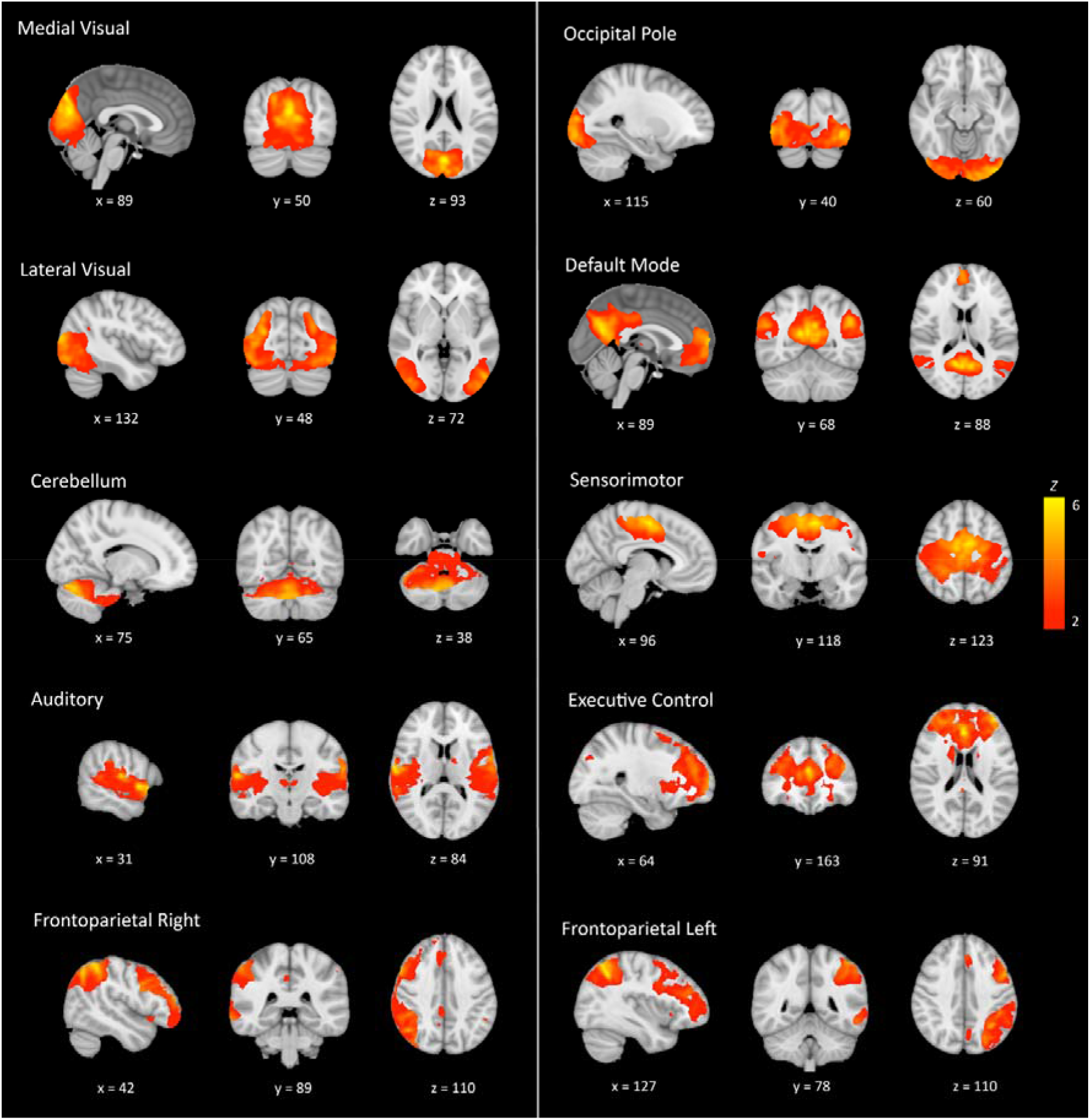
Resting state networks derived from group ICA. Group ICA was performed using ICA-AROMA cleaned components of control subject with the component most closely resembling previously described networks (26) selected to represent that network. Displayed are components converted to a Z-score overlaid on the MNI152 standard brain. X, y, and z values refer to the voxel location of the image. ICA = independent component analysis.

### Regions of the executive network showed significantly decreased activity in limb-onset ALS patients

Comparisons between groups showed significantly decreased activity within a region within the executive control network (EXN) in limb-onset ALS subjects (P < 0.05 after TFCE; Figure 2). All other networks were insignificantly different, however the occipital pole and frontoparietal networks were nearing significance (Supplementary Table 1).

**Figure 2.**
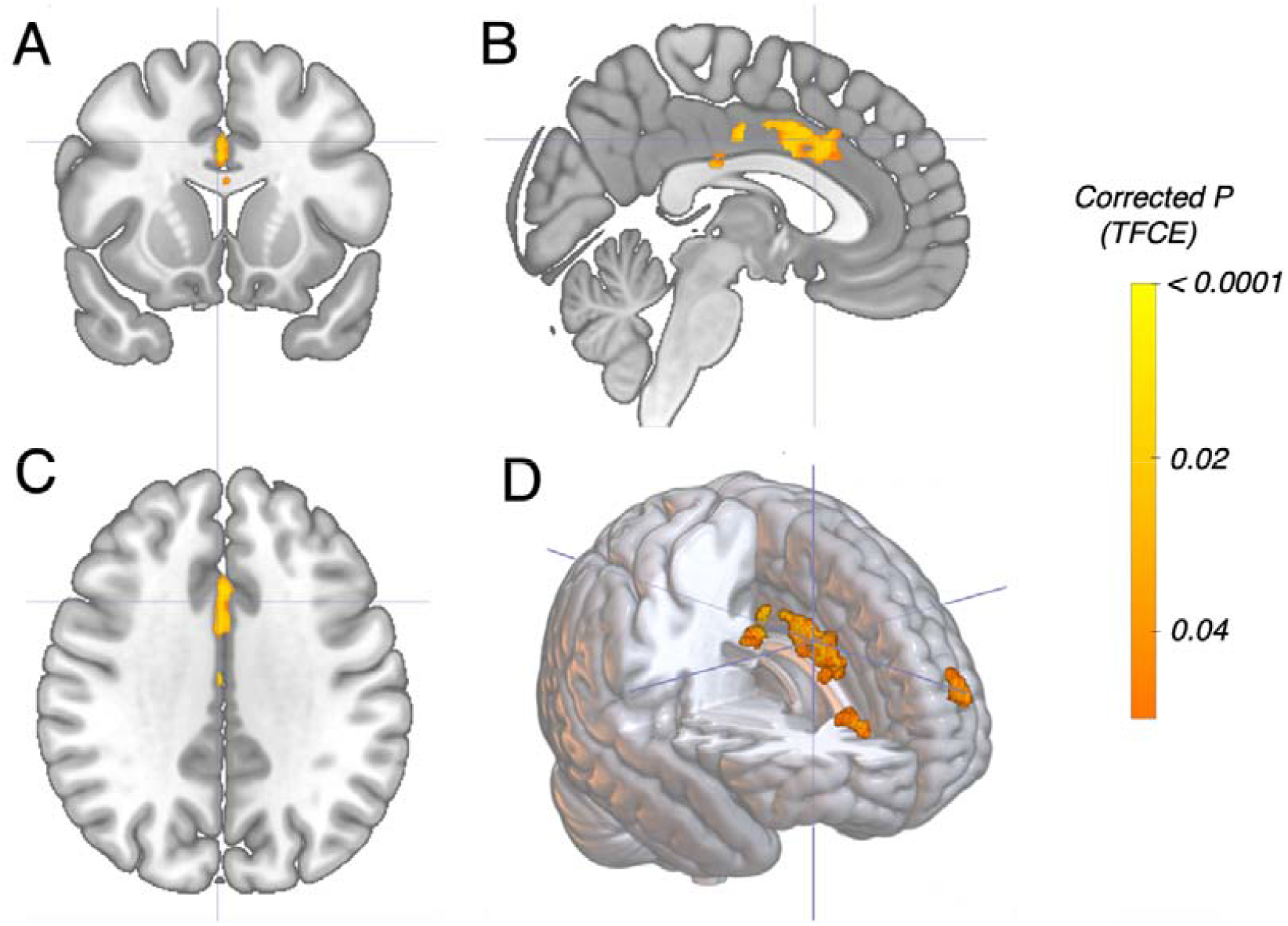
Regions of the EXN showing significantly decreased activity in ALS patients compared to controls. In orange are significant P values after TFCE (α = 0.05) overlaid on (A) axial, (B) coronal, (C) sagittal, and (D) 3-d render views of the MNI152 standard brain. EXN = executive control network, TFCE = threshold-free cluster enhancement.

Overall, there were 2,671 voxels within this EXN cluster, which localized primarily to the anterior cingulate gyrus (2,134 voxels – 79.90% of total), with smaller clusters in the frontal pole (533 – 19.96%) and paracingulate gyrus (4 – 0.15%) (Table 2).

**Table 2.** Localization of significant EXN cluster. The number of voxels belonging to anatomical regions of the significant EXN cluster were calculated and displayed as a total percentage of the cluster.

| Anatomical Region | Number of voxels | Percentage of cluster (%) |
| --- | --- | --- |
| Anterior Cingulate Gyrus | 2,134 | 79.90 |
| Frontal Pole | 533 | 19.96 |
| Paracingulate Gyrus | 4 | 0.15 |
| Total | 2,671 | N/A |

### Accurate SVM classification of limb-onset ALS patients using activity within EXN cluster

Mean ß-coefficients from individual spatial maps were calculated for each of the RSNs as well as the significant EXN cluster, which were used to train a SVM classifier evaluated with ten-fold cross-validation. The SVM trained using mean EXN cluster ß-coefficients showed excellent performance on both the training (AUC = 0.93) and validation (AUC =0.93) datasets (Figure 3A & B, respectively). The optimal threshold of 0.65 which returned a sensitivity of 86% with 87% specificity (Supplementary Table 2). Excluding the entire EXN on the training data with an AUC of 0.71, all other networks displayed near random performance on both datasets (AUC range: 0.34 – 0.62).

**Figure 3.**
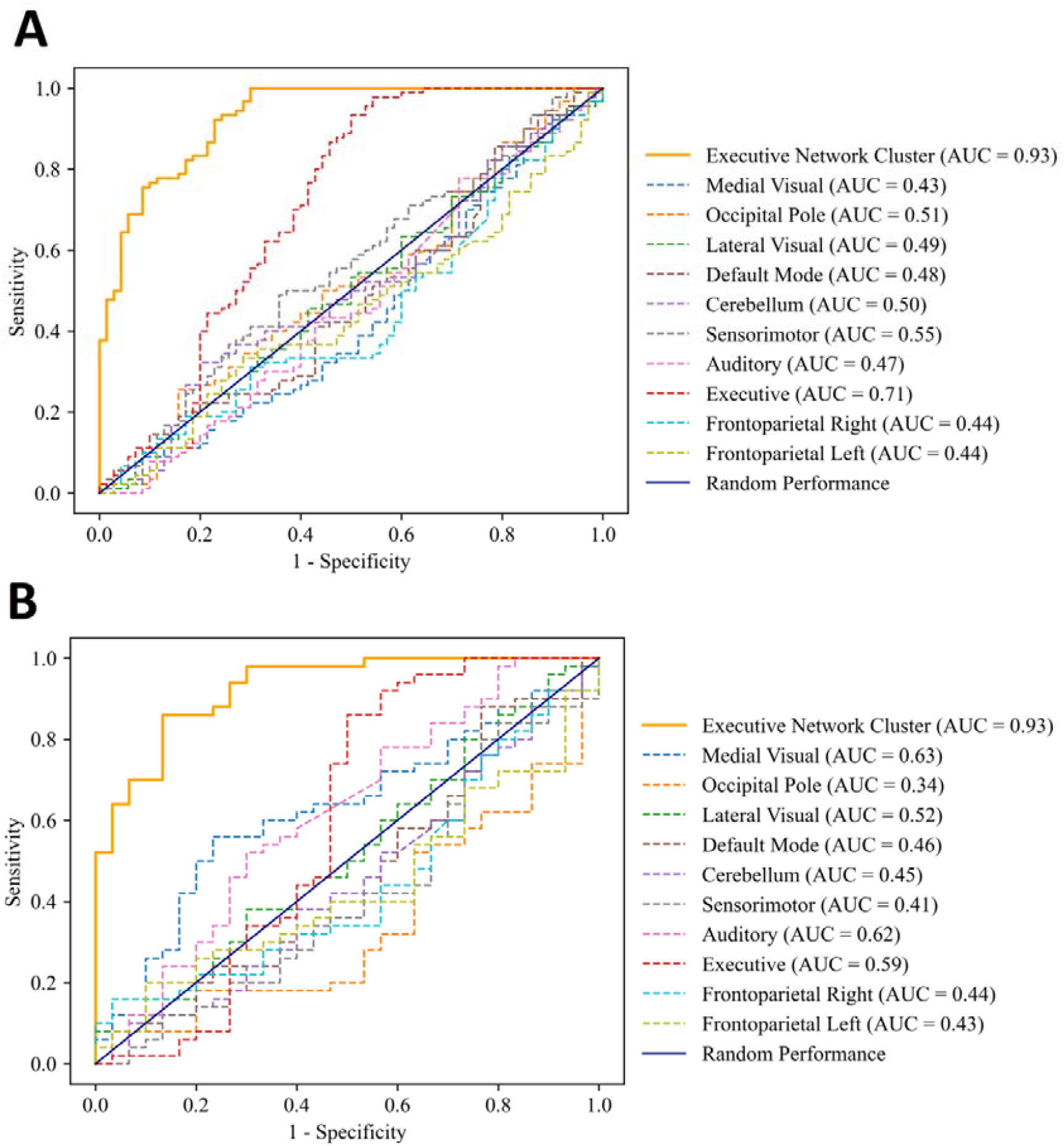
Accurate SVM classification of limb-onset ALS patients using EXN cluster. Shown are ROC curves for SVMs trained using β-coefficients of each RSN (dotted lines) and EXN cluster (orange solid line) on (A) training and (B) validation datasets. Solid navy line represents random performance. The EXN cluster SVM excellent performance on both the training (AUC = 0.93) and validation (AUC = 0.93) datasets. SVM = support vector machine, ROC = receiver operator characteristic, AUC = area-under-curve.

## Discussion

Amyotrophic lateral sclerosis (ALS) is highly heterogeneous, and its similar presentation to other neurological disorders creates diagnostic challenges. In other neurological disorders, multi-modal classifiers have shown efficacy, and a similar approach could be employed in ALS, highlighting the importance of discovering neuroimaging biomarkers with classification potential.

This study assessed resting-state network (RSN) alterations in a purely limb-onset ALS cohort. Mean β-coefficients of a cluster within the executive control network (EXN), significantly decreased in limb-onset ALS patients, was used to train a support vector machine (SVM) classifier with 86% sensitivity, 87% specificity, and area-under-curve (AUC) of 0.93 on the validation dataset at the optimal threshold. These results support our hypothesis that functionally altered regions in limb-onset ALS will be able to develop an SVM with greater than 80% sensitivity and specificity. This improves on past studies utilizing independent component analysis (ICA) that developed a SVM with an AUC of 0.72 (7) and builds on the body of literature showing classification potential using structural, functional, diffusion, and spinal cord MRI (4–8), and quantitative susceptibility mapping (QSM) metrics (9,10). Altogether, this study provides another metric that could be included in multi-modal classification algorithms in future. Given functional alterations are thought to precede structural changes and symptom development (12), our findings present an opportunity for early detection.

### Decreased activity in executive control network of limb-onset ALS patients

Between-group analysis showed a region of the EXN significantly decreased in limb-onset ALS patients localizing to the anterior cingulate gyrus (ACG), frontal pole, and paracingulate gyrus. It is worth noting that differences in the frontoparietal networks (FTPNs) and occipital pole network were nearing significance, which may not have been detected due to sample size, which was less than half the recommended size to detect moderate to strong effects in neuroimaging studies (31,32). This trend was mentioned as several studies have shown decreased activity in temporo-occipital regions of several networks – including the FTPN and visual networks – in heavily (>80%) limb-onset ALS cohorts (13,33,34). Based on this observation, it should be explored if temporo-occipital decreases are a hallmark of this subtype.

Strengthening the involvement of these regions in this subtype are longitudinal studies in heavily limb-onset ALS cohorts which describe progressive increases over time (35–37). Given the suggestion of decreased activity compared to controls (13,33,34), it is unclear whether these progressive increases are a compensatory mechanism following these initial decreases, or reflect interneuron degeneration causing decreased inhibition, which has been demonstrated in mouse models (38,39) and post-mortem brain tissue (40). The neurodegenerative mechanisms of ALS and its implications on functional activity, particularly within temporo-occipital regions of the brain, should be a focus of future work.

### Limitations

This study had several limitations, namely sample size, which may contribute to the inability to find significant effects, given studies investigating effect-sizes suggest each group should have 40 subjects to detect strong effects, and over 80 to detect moderate effects (31,32). Furthermore, an increased sample size would increase validation dataset size for the SVM. Although this was mitigated through cross-validation, increased validation cohorts would improve the reliability of our findings.

Secondly, our cohort was very skewed, with 12 of the 14 limb-onset ALS patients being male. As RSN sex-differences have been reported in ALS (41) this is both a strength, as it improves the specificity of our cohort, but also a limitation, as it decreases the translatability of findings to females. Finally, our cerebellum RSN map showed poor similarity to previously described RSNs (26) and this may affect detecting effects in this region.

## Conclusion

This study found a region of the executive control network (EXN) significantly decreased in limb-onset ALS patients compared to controls, localized predominantly to the anterior cingulate gyrus (ACG). Furthermore, activity within this region was used as input for support vector machine classifier, which showed excellent sensitivity and specificity in classifying patients. Use of activity within regions of the EXN, and the ACG specifically, could be incorporated with other neuroimaging metrics into multi-modal classifiers to address the diagnostic gap in ALS.

## Data Availability

All data produced in the present study are available upon reasonable request to the authors

## Acknowledgements

We would like to thank all the volunteers who participated in this study. We are also grateful to the radiographers at Monash Biomedical Imaging for their assistance with MRI data collection and our nurse research assistants Ruth Krasniqi and Anna Smith. Funding for this project was obtained through the Monash University Strategic Grant Scheme.

