## Supplementary Table 2 for "Accurate support vector machine identification of limb-onset amyotrophic lateral sclerosis using resting-state activity of regions within executive control network"

**Supplementary Table 2. Performance of SVM trained on EXN cluster activity.** *Shown is* *validation dataset* *sensitivity and specificity of the SVM trained using EXN ROI activity across a range of thresholds. The optimal threshold of 0.65 returned a sensitivity of 86% and specificity of 87%.*

| **Threshold** | **Sensitivity** | **Specificity** |
| --- | --- | --- |
| 0.78 | 0.02 | 1.00 |
| 0.69 | 0.52 | 0.97 |
| 0.68 | 0.64 | 0.93 |
| 0.67 | 0.70 | 0.87 |
| 0.65 | 0.86 | 0.87 |
| 0.64 | 0.88 | 0.77 |
| 0.63 | 0.94 | 0.73 |
| 0.62 | 0.98 | 0.70 |
| 0.52 | 0.98 | 0.47 |
| 0.49 | 1.00 | 0.47 |
| 0.05 | 1.00 | 0.00 |
