## Supplementary Table 1 for "Accurate support vector machine identification of limb-onset amyotrophic lateral sclerosis using resting-state activity of regions within executive control network"

**Supplementary Table 1. P-values for between-group contrasts.** *Shown are P-values after TFCE correction in all RSNs. TFCE = threshold-free cluster enhancement, RSNs = resting-state network.*

| **Resting-state network** | **ALS > Control** | **Control > ALS** |
| --- | --- | --- |
| Medial Visual | 0.218 | 0.886 |
| Occipital Pole | 0.601 | 0.079 |
| Lateral Visual | 0.546 | 0.404 |
| Default Mode | 0.211 | 0.744 |
| Cerebellum | 0.166 | 0.441 |
| Sensorimotor | 0.324 | 0.207 |
| Auditory | 0.316 | 0.772 |
| Executive Control | 0.864 | 0.011* |
| Frontoparietal Right | 0.543 | 0.091 |
| Frontoparietal Left | 0.654 | 0.134 |

**Supplementary Table 2. Performance of SVM trained on EXN cluster activity.** *Shown is* *validation dataset* *sensitivity and specificity of the SVM trained using EXN ROI activity across a range of thresholds. The optimal threshold of 0.65 returned a sensitivity of 86% and specificity of 87%.*

| **Threshold** | **Sensitivity** | **Specificity** |
| --- | --- | --- |
| 0.78 | 0.02 | 1.00 |
| 0.69 | 0.52 | 0.97 |
| 0.68 | 0.64 | 0.93 |
| 0.67 | 0.70 | 0.87 |
| 0.65 | 0.86 | 0.87 |
| 0.64 | 0.88 | 0.77 |
| 0.63 | 0.94 | 0.73 |
| 0.62 | 0.98 | 0.70 |
| 0.52 | 0.98 | 0.47 |
| 0.49 | 1.00 | 0.47 |
| 0.05 | 1.00 | 0.00 |
